# Access to primary healthcare during lockdown measures for COVID-19 in rural South Africa: a longitudinal cohort study

**DOI:** 10.1101/2020.05.15.20103226

**Authors:** Mark J. Siedner, John D. Kraemer, Mark J. Meyer, Guy Harling, Thobeka Mngomezulu, Patrick Gabela, Siphephelo Dlamini, Dickman Gareta, Nomathamsanqa Majozi, Nothando Ngwenya, Janet Seeley, Emily Wong, Collins Iwuji, Maryam Shahmanesh, Willem Hanekom, Kobus Herbst

**Affiliations:** Massachusetts General Hospital; Georgetown University; University College London; Africa Health Research Institute; London School of Hygiene and Tropical Medicine; Brighton and Sussex Medical School

**Author notes:** Corresponding Author Mark J. Siedner Africa Health Research Institute KwaZulu-Natal, South Africa.

**Keywords:** COVID-19, South Africa, Primary Care, Health Systems Resilience, Health and Demographic Surveillance System

## Abstract

**Objectives:** Public health interventions designed to interrupt COVID-19 transmission could have deleterious impacts on primary healthcare access. We sought to identify whether implementation of the nationwide lockdown (shelter-in-place) order in South Africa affected ambulatory clinic visitation in rural Kwa-Zulu Natal (KZN).

**Design:** Prospective, longitudinal cohort study

**Setting:** Data were analyzed from the Africa Health Research Institute Health and Demographic Surveillance System, which includes prospective data capture of clinic visits at eleven primary healthcare clinics in northern KwaZulu-Natal

**Participants:** A total of 36,291 individuals made 55,545 clinic visits during the observation period.

**Exposure of Interest:** We conducted an interrupted time series analysis with regression discontinuity methods to estimate changes in outpatient clinic visitation from 60 days before through 35 days after the lockdown period.

**Outcome Measures:** Daily clinic visitation at ambulatory clinics. In stratified analyses we assessed visitation for the following sub-categories: child health, perinatal care and family planning, HIV services, noncommunicable diseases, and by age and sex strata.

**Results:** We found no change in total clinic visits/clinic/day from prior to and during the lockdown (–6.9 visits/clinic/day, 95%CI –17.4, 3.7) or trends in clinic visitation over time during the lockdown period (–0.2, 95%CI –3.4, 3.1). We did detect a reduction in child healthcare visits at the lockdown (–7.2 visits/clinic/day, 95%CI –9.2, –5.3), which was seen in both children <1 and children 1–5. In contrast, we found a significant increase in HIV visits immediately after the lockdown (8.4 visits/clinic/day, 95%CI 2.4, 14.4). No other differences in clinic visitation were found for perinatal care and family planning, non-communicable diseases, or among adult men and women.

**Conclusions:** In rural KZN, the ambulatory healthcare system was largely resilient during the national-wide lockdown order. A major exception was child healthcare visitation, which declined immediately after the lockdown but began to normalize in the weeks thereafter. Future work should explore efforts to decentralize chronic care for high-risk populations and whether catch-up vaccination programs might be required in the wake of these findings.

**What is already known on this topic?:**

- Prior disease epidemics have created severe interruptions in access to primary care in sub-Saharan Africa, resulting in increased child and maternal mortality
- Data from resource-rich settings and modelling studies have suggested the COVID-19 epidemic and non-pharmacologic measures implemented in response could similarly result in substantial barriers to primary health care access in the region
- We leveraged a clinical information system in rural KwaZulu-Natal to empirically assess the effect of the COVID-19 epidemic and a nationwide lockdown in South Africa on access to primary care

**What this study adds?:**

- Access to primary healthcare was largely maintained during the most stringent period of the COVID-19 lockdown in South Africa, with the exception of a temporary drop in child health visits
- Creative solutions are needed for sustaining child vaccination programs, and protecting high-risk individuals from risk of nosocomial transmission in resource-limited settings

## Introduction

COVID-19 was declared a global pandemic by the World Health Organization on 11^th^ March 2020, and it has spared no region of the world. Thus far, the greatest numbers of cases have been reported in Asia, Europe and North America.^1^ Limited testing and surveillance capabilities make it difficult to assess how widely the pandemic has spread in low-resource settings. But such regions are believed to be at particular risk of severe epidemics, due to over-crowding, lower access to clean water and sanitation services, and inherent shortages in health system infrastructure for detection and management of disease.^2–9^

In response, most nations in sub-Saharan Africa have implemented non-pharmacologic interventions to attempt to prevent large scale epidemics. These measures, which include restrictions on large gatherings, work and school attendance, travel, and in their most stringent forms, shelter-in-place orders, are believed to reduce disease transmission.^10–13^ However, instituting these measures is also associated with deleterious economic, social, and health impacts.^14–16^ Some have hypothesized that non-pharmaceutical interventions might be less effective in settings with large informal economies and limited ability to respond to increases in cases of severe disease,^17^ and that their risks might outweigh their benefits.^18^

Of particular concern is how social fear and reduced access to basic public health services might impact morbidity and mortality for non-COVID health conditions, including perinatal and childcare, chronic communicable and non-communicable disease, and emergency care services. Modeling studies have suggested that even modest reductions in child healthcare access could result in 100,000s of additional deaths in low and middle-income countries.^19^ Interruptions in basic healthcare access during recent Ebola epidemics were associated with increases in morbidity and mortality.^20 21^ Yet, whether such effects will be seen during the COVID-19 epidemic is not known.

On 27^th^ March, 2020, South Africa instituted a nationwide shelter-in-place order, termed in South Africa as a national Level 5 lockdown (with levels ranging from 1 to 5, and 5 being the most stringent level of social distancing).^22^ The order included closure of schools and all non-essential business, restrictions on public transport, and restrictions on movement. The healthcare sector was deemed an essential service, and no restrictions were placed on access to or delivery of healthcare services. We sought to assess the impact of the lockdown order in response to the COVID-19 epidemic in South Africa on access to basic healthcare services. We analysed data on clinic visitation at 11 ambulatory public health clinics in northern KwaZulu-Natal, collected routinely as part of a demographic health and surveillance system (HDSS) by the Africa Health Research Institute (AHRI). We hypothesized that there would be immediate and substantial reductions in clinic visitation after the institution of the lockdown measure, and that this would pertain to routine clinical care such as immunizations, perinatal care, and chronic disease management.

## Methods

### Study Setting

This analysis was conducted using data collected by the AHRI HDSS in the uMkhanyakude district of the KwaZulu-Natal province. The HDSS comprises a complete census across a geographic area of approximately 850 km^2^; it is a rural region with a single peri-urban centre, KwaMsane, a town of approximately 30,000 residents. The region ranks among the lowest nationwide in terms of health indicators and socioeconomic status.^23^ Approximately 1 in 5 adult men and 2 in 5 adult women are living with HIV.^24^ Tuberculosis incidence is among the highest in the world, and above the national average of 577 per 100,000 individuals when last measured in 2015.^25^

### Data Collection

Since 2000 AHRI has collected data on births, deaths, migrations through thrice annual data collection encounters across a catchment area of 20,000 households (over 100,000 resident individuals).^23^ In 2017, AHRI began placing clinic research assistants at each of the 11 government-run public health clinics in the area. These research staff operate in partnership with the Department of Health, but outside of the standard Health Management Information System (HMIS). For each person who presents to clinic, they collect demographic information and the self-reported reason(s) for the clinical visit. Data is electronically captured and linked by a unifying identification code to the HDSS using a Clinic-Link data syncing system developed by AHRI. AHRI holds memoranda of understanding with the Provincial and District Department of Health that permit extraction of health record data from primary care and hospital sites for linkage to the household surveillance dataset.

### Study Design

We conducted an interrupted time series analysis to estimate changes in clinic visitation in rural KwaZulu-Natal from before to after the national lockdown implementation on 27^th^ March 2020. To do so we fit mixed effects linear regression models by restricted maximum likelihood with daily clinic visits as the primary outcome of interest. Our primary exposure of interest was time period, divided into the pre-lockdown period 60-days prior to the lockdown date, and the lockdown-period, starting 27^th^ March 2020 through 30^th^ April 2020, the last day before South Africa transitioned to a Level 4 lockdown. We estimated the change in clinic mean visits per clinic at the date of the lockdown and the change in mean visits per clinic over time after the lockdown using regression discontinuity methods,^26^ which allows us to estimate both the immediate impact of the lockdown and trend in visit daily after it went into place. We included a fixed effect for day of the week and a random effect for clinic. We excluded weekends because most of the ambulatory clinics observed do not operate on weekends. We excluded dates from observation when AHRI staff members who perform data capture for the Clinic-link system were not working, including national holidays and staff trainings.

Our primary outcome of interest was the number of clinic visits for any reason per facility per day. In secondary analyses, we stratified models by visit type restricted to: 1) child health visits (immunizations and growth monitoring); 2) antenatal care, postnatal care, and family planning; 3) HIV services (including antiretroviral therapy initiation, antiretroviral therapy continuation, and chronic care medical dispensing program visits); and 4) chronic care of non-communicable diseases (hypertension and diabetes). Clinic visit for more than one reason were treated as visits for both conditions. We also conducted stratified analyses by age category (<1, 1–5, 6–19, 20–45, and > 45 years old) and by women and men aged 15 years or older.

To test for robustness to model assumptions, we conducted four sensitivity analyses: 1) we added random slopes by time to the main linear mixed effects model to account for possible temporal autocorrelation and tested this model against the main model with a likelihood ratio test; we fit 2) linear and 3) Poisson generalized estimating equation models clustered by facility; and 4) we added a quadratic term for time during the post-lockdown period to assess for a non-linear relationship between time and clinic visitation.

Finally, we conducted an additional sensitivity analysis to assess for the occurrence of in-migration into the HDSS catchment area during the lockdown period, which would potentially bias clinic visitation frequency upwards. To do so, we calculated annual visitation frequency at the 11 area clinics for each individual in the dataset for the year prior to the lockdown. We then compared the median number of annual visits per individual in the pre- and post-lockdown periods, and the number of individuals with exactly one visit in the past year in the two periods. If a significant in-migration did occur during the lockdown period, we would expect that the median number of annual visits per individual would decrease during the lockdown, whereas the number of individuals with one visit in the past 12 months would increase.

### Patient and Public Involvement

This protocol was reviewed and approved by the AHRI Community Advisory Board, who contributed input on the study design and collection measures. Results of studies from the HDSS project are routinely shared with the community through public communications and road shows conducted by the AHRI Public Engagement Department. Final, all study protocols are reviewed and approved by the District and Provincial Department of Health, and AHRI holds memoranda of understanding with the Provincial and District Departments of Health that outline methods of extraction of health record data from primary care sites for linkage to the household surveillance dataset.

### Ethical Approval

The protocol was reviewed and approved by the University of KwaZulu-Natal Biomedical Research Ethics Committee under reference BE290/16 and the KwaZulu Department of Health Research Committee.

## Results

A total of 36,291 individuals made 55,545 clinic visits between 27^th^ January – 29th April 2020 at the 11 area clinics (**Table 1**). Women and girls accounted for 70% (n = 25,393) of visits. Approximately 9% of visits were made by individuals less than 1 (n = 3,124), 1–5 (n = 3,125), and 6–19 years old (n = 3,175), respectively; whereas those 20–45 years accounted for 47% (n = 17,226) and those over 46 the remaining 27% of visits (n = 9,642). The most common reason for a clinic visit was ART follow-up care, comprising 40% of all visits (n = 22,243), followed by visits for minor ailments (20%, n = 11,049), child health (n = 6,194, 11%) and hypertension (n = 5,790, n = 10%).

**Table 1.** Ambulatory clinic visits at 11 region clinics in rural KwaZulu Natal during 27 January 2020 – 30 April 2020 by sex and age and clinic visit type

|  | **Total\***<br>**n (%)** | **Male**<br>**n (%)** | **Female**<br>**n (%)** | **<1 year**<br>**n (%)** | **1-5 years**<br>**n (%)** | **6-19 years**<br>**n (%)** | **20-45 years**<br>**n (%)** | **>45 years**<br>**n (%)** |
| --- | --- | --- | --- | --- | --- | --- | --- | --- |
| Total visits | 55,545<br>(100%) | 16,082<br>(29.0%) | 39,444<br>(71.0%) | 4,987<br>(9.0%) | 3,914<br>(7.1%) | 4,530<br>(8.2%) | 26,196<br>(47.2%) | 15,918<br>(28.7%) |
| Child health | 6194<br>(11.2%) | 3081<br>(49.7%) | 3104<br>(50.1%) | 4270<br>(68.9%) | 1786<br>(28.8%) | 103<br>(1.7%) | 29<br>(0.5%) | 6<br>(0.1%) |
| PNC and FP<sup>a</sup> | 4,634<br>(8.3%) | 6<br>(0.0%) | 4,628<br>(11.7%) | 6<br>(0.1%) | 1<br>(0.0%) | 746<br>(16.5%) | 3,863<br>(14.8%) | 18<br>(0.1%) |
| HIV visit<sup>b</sup> | 25,550<br>(46.0%) | 6,791<br>(43.3%) | 18,755<br>(47.6%) | 25<br>(0.5%) | 131<br>(3.4%) | 1,543<br>(34.1%) | 16,265<br>(62.1%) | 7,586<br>(47.7%) |
| Chronic care<sup>c</sup> | 6,290<br>(11.3%) | 1,355<br>(8.4%) | 4,935<br>(12.5%) | 1<br>(0%) | 0 | 4<br>(0.1%) | 411<br>(1.6%) | 5,874<br>(36.9%) |
| Child health | 6194<br>(11.2%) | 3081<br>(49.7%) | 3104<br>(50.1%) | 4270<br>(68.9%) | 1786<br>(28.8%) | 103<br>(1.7%) | 29<br>(0.5%) | 6<br>(0.1%) |
| Minor ailment | 12,751<br>(23.0%) | 4,220<br>(26.2%) | 8,525<br>(21.6%) | 755<br>(15.1%) | 2,043<br>(52.2%) | 1,969<br>(43.5%) | 5,548<br>(21.2%) | 2,436<br>(15.3%) |
| All other visits | 4,637<br>(8.4%) | 913<br>(5.8%) | 3,706<br>(9.4%) | 9<br>(0.2%) | 34<br>(0.9%) | 809<br>(17.9%) | 3,158<br>(12.1%) | 627<br>(3.9%) |

\*Visit types are not mutually exclusive
<sup>a</sup>PNC and FP: Perinatal care and family planning; visits for, antenatal care, prenatal care, and/or family planning
<sup>b</sup>HIV visits: visits for HIV testing, antiretroviral therapy initiation, antiretroviral therapy continuation, or pharmacy pick-up
<sup>c</sup>Chronic care: clinical visits for hypertension and/or diabetes

There was an average of 89.2 (95%CI 65.5, 112.9) clinic visits per day per clinic in the pre-lockdown period, with a non-significant drop immediately following the lockdown (−6.9 visits/clinic/day, 95%CI –17.4, 3.7), and no significant change in trend from the pre- to post-lockdown period (**Table 2**, Figure 1). Child health visits decreased by over 50% from before to immediately after the lockdown (from 11.8 to 4.5 visits/day/clinic, mean change of –7.2 visits, 95%CI –9.2, –5.3) but then partially rebound in the post-lockdown period (+1.1 visit/clinic/day with each passing week [95%CI 0.5, 1.7]). In contrast to child health visits, clinical visits for HIV services increased by approximately 20% immediately after the start of the lockdown(from 37.7 to 46.1, for an increase of 8.4 visits/clinic/day [95%CI 2.4, 14.4]). Like child health visits, this initial increase was followed by trend in the opposite direction, (–1.5 visit/clinic/day with each passing week, 95%CI –3.4, 0.3).

**Table 2.**
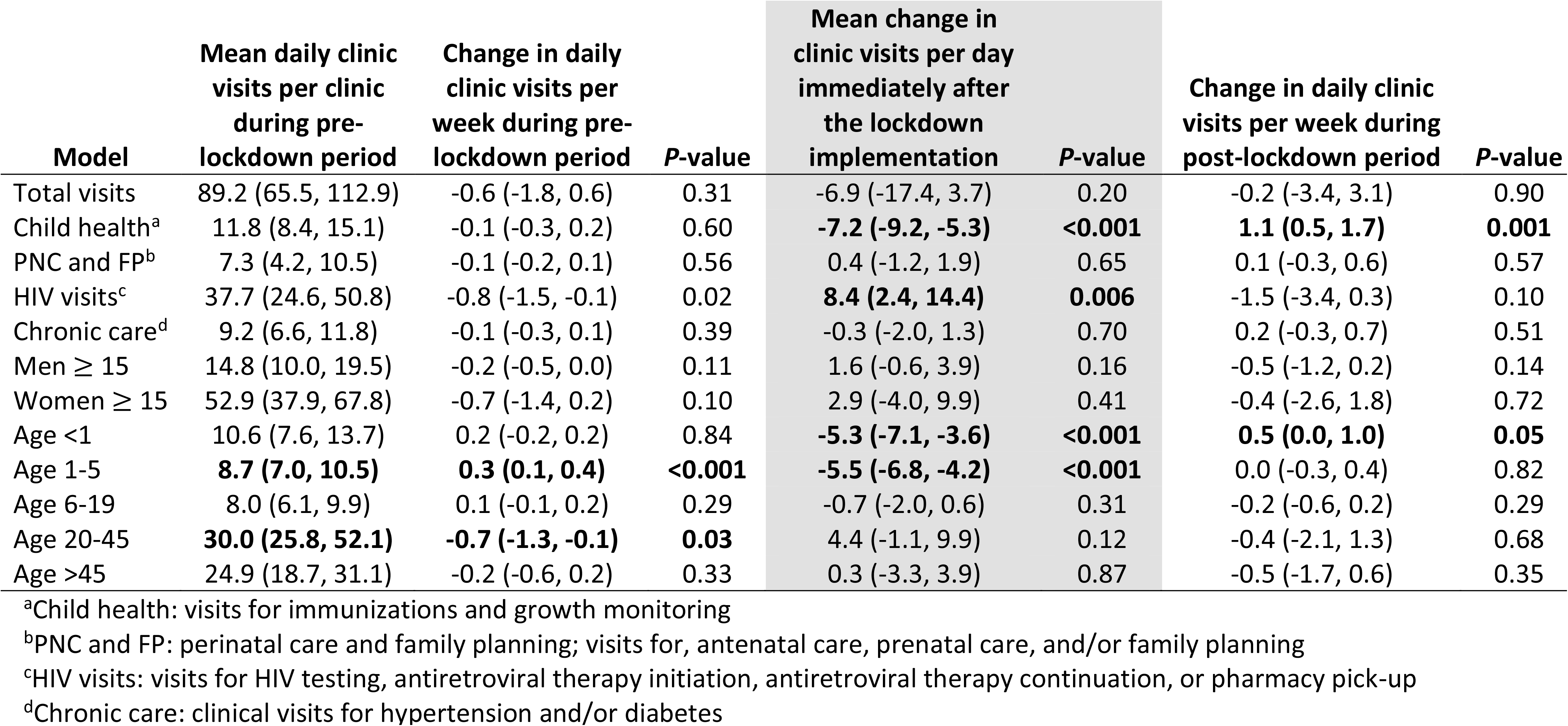
Mixed effects regression model results demonstrating mean clinic visits overall, by clinic type and demographic strata, in the pre- and post-lockdown period in uMkhanyakude District, KwaZulu-Natal South Africa.

| Model | Mean daily clinic visits per clinic during pre-lockdown period | Change in daily clinic visits per week during pre-lockdown period | P-value | Mean change in clinic visits per day immediately after the lockdown implementation | P-value | Change in daily clinic visits per week during post-lockdown period | P-value |
| --- | --- | --- | --- | --- | --- | --- | --- |
| Total visits | 89.2 (65.5, 112.9) | -0.6 (-1.8, 0.6) | 0.31 | -6.9 (-17.4, 3.7) | 0.20 | -0.2 (-3.4, 3.1) | 0.90 |
| Child health<sup>a</sup> | 11.8 (8.4, 15.1) | -0.1 (-0.3, 0.2) | 0.60 | **-7.2 (-9.2, -5.3)** | **<0.001** | **1.1 (0.5, 1.7)** | **0.001** |
| PNC and FP<sup>b</sup> | 7.3 (4.2, 10.5) | -0.1 (-0.2, 0.1) | 0.56 | 0.4 (-1.2, 1.9) | 0.65 | 0.1 (-0.3, 0.6) | 0.57 |
| HIV visits<sup>c</sup> | 37.7 (24.6, 50.8) | -0.8 (-1.5, -0.1) | 0.02 | **8.4 (2.4, 14.4)** | **0.006** | -1.5 (-3.4, 0.3) | 0.10 |
| Chronic care<sup>d</sup> | 9.2 (6.6, 11.8) | -0.1 (-0.3, 0.1) | 0.39 | -0.3 (-2.0, 1.3) | 0.70 | 0.2 (-0.3, 0.7) | 0.51 |
| Men ≥ 15 | 14.8 (10.0, 19.5) | -0.2 (-0.5, 0.0) | 0.11 | 1.6 (-0.6, 3.9) | 0.16 | -0.5 (-1.2, 0.2) | 0.14 |
| Women ≥ 15 | 52.9 (37.9, 67.8) | -0.7 (-1.4, 0.2) | 0.10 | 2.9 (-4.0, 9.9) | 0.41 | -0.4 (-2.6, 1.8) | 0.72 |
| Age <1 | 10.6 (7.6, 13.7) | 0.2 (-0.2, 0.2) | 0.84 | **-5.3 (-7.1, -3.6)** | **<0.001** | **0.5 (0.0, 1.0)** | **0.05** |
| Age 1-5 | **8.7 (7.0, 10.5)** | **0.3 (0.1, 0.4)** | **<0.001** | **-5.5 (-6.8, -4.2)** | **<0.001** | 0.0 (-0.3, 0.4) | 0.82 |
| Age 6-19 | 8.0 (6.1, 9.9) | 0.1 (-0.1, 0.2) | 0.29 | -0.7 (-2.0, 0.6) | 0.31 | -0.2 (-0.6, 0.2) | 0.29 |
| Age 20-45 | **30.0 (25.8, 52.1)** | **-0.7 (-1.3, -0.1)** | **0.03** | 4.4 (-1.1, 9.9) | 0.12 | -0.4 (-2.1, 1.3) | 0.68 |
| Age >45 | 24.9 (18.7, 31.1) | -0.2 (-0.6, 0.2) | 0.33 | 0.3 (-3.3, 3.9) | 0.87 | -0.5 (-1.7, 0.6) | 0.35 |

<sup>a</sup>Child health: visits for immunizations and growth monitoring
<sup>b</sup>PNC and FP: perinatal care and family planning; visits for, antenatal care, prenatal care, and/or family planning
<sup>c</sup>HIV visits: visits for HIV testing, antiretroviral therapy initiation, antiretroviral therapy continuation, or pharmacy pick-up
<sup>d</sup>Chronic care: clinical visits for hypertension and/or diabetes

**Figure 1.**
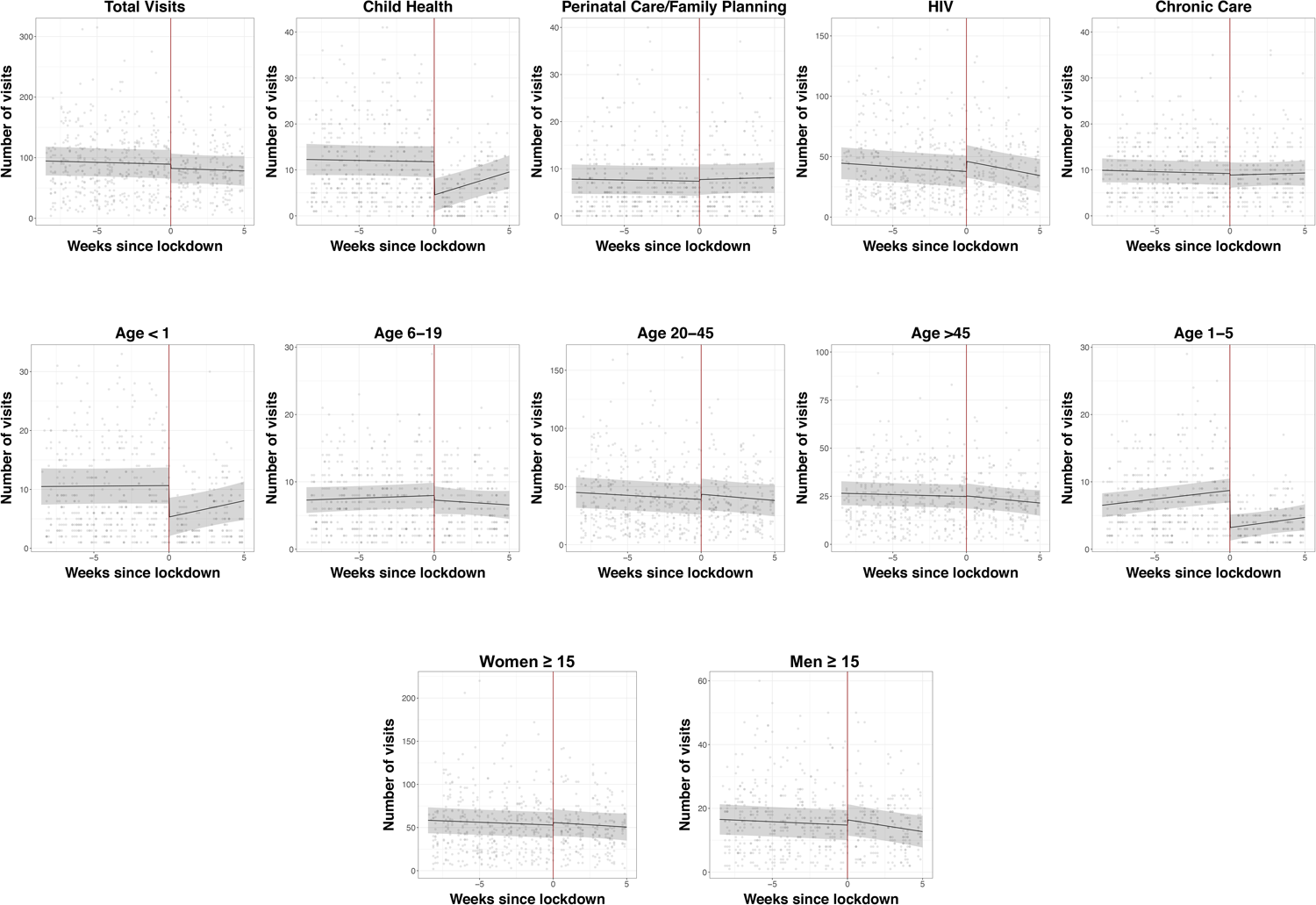
Ambulatory clinic visitation before and after the nationwide lockdown in South Africa at eleven outpatient clinics in rural uMkhanyakude District, KwaZulu-Natal South Africa. Scatterplot represents mean clinic visitation at each clinic on weekdays during the observation period. The black fit line represents the mean visitation across all clinics estimated by post-regression margins from a linear regression model, with a regression discontinuity coefficient at the date of the lockdown (27^th^ March 2020, red line). Gray bars represent 95% confidence intervals.

In age-stratified analyses, we observed significant reductions of more than 50% for children under 1 (10.6 to 5.3, mean decrease of –5.3 visits, 95%CI –7.1, –3.6) and 1–5 years old (8.7 to 3.2, mean decrease of –5.5 visits, 95%CI –6.8, –4.2), with a partial rebound of infant visits(change in trend = 0.5 visits per week, 95%CI 0.0, 1.0) but not in visitation frequency by 1–5 year old children. We did not find changes in clinic visitation for chronic non-communicable diseases or perinatal and family planning visits, or changes in clinic visitation by men or women 15 years or older.

Results were robust to modelling assumptions in the sensitivity analyses (**Table 3**). The addition of random slopes in the primary model was not associated with an improvement in model fitness (likelihood ratio test chi-squared 3.26, *P*–value = 0.07. We also did not find evidence of a non-linear relationship between time and clinic visitation in the primary model (*P* = 0.50 for the quadratic term).

**Supplemental Table 3.** Sensitivity analyses, demonstrating results of the main regression model and alternate models including a mixed effects regression model with random slopes, and linear and Poisson generalized estimating equations models.

| Model | Mean daily clinic visits per clinic during pre-lockdown period | Change in daily clinic visits per week during pre-lockdown period | P-value | Mean change in clinic visits per day immediately after the lockdown implementation |  | Change in daily clinic visits per week during post-lockdown period | P-value |
| --- | --- | --- | --- | --- | --- | --- | --- |
| Primary model | 89.2 (65.5, 112.9) | -0.6 (-1.8, 0.6) | 0.31 | -6.9 (-17.4, 3.7) | 0.20 | -0.2 (-3.4, 3.1) | 0.90 |
| Random slopes | 89.3 (66.0, 112.5) | -0.6 (-1.9, 0.7) | 0.38 | -6.9 (-17.4, 3.5) | 0.19 | -0.2 (-3.4, 3.1) | 0.89 |
| Linear GEE | 89.2 (66.9, 111.5) | -0.6 (-1.8, 0.6) | 0.31 | -6.9 (-17.5, 3.8) | 0.21 | -0.2 (-3.5, 3.1) | 0.90 |
| Poisson GEE<sup>b</sup> | 89.2 (84.8, 93.8) | 1.0 (0.9, 1.0) | <0.001 | 0.9 (0.9, 0.9) | <0.001 | 1.0 (0.9, 1.0) | 0.30 |

GEE: generalized estimating equations
<sup>a</sup>Random slopes model is a linear mixed effects regression model with random intercept by clinic and random slope by time
<sup>b</sup>Poisson GEE results are presented as exponentiated coefficients, so they represent proportional rather than absolute changes in counts.

In our final sensitivity analysis, we did not detect evidence of meaningful in-migration. The median number of annual visits per individual attending the clinic did not differ between the pre- (median 5, interquartile range [IQR] 2–7) and post-lockdown periods (median 5, IQR 2–7), *P* = 0.67. This pattern was similar among people attending clinic for HIV-specific visits (median 5 [IQR 3–7] vs median 5 [IQR 3–7], *P* = 0.36. The number of people with exactly one visit in the past year also did not meaningfully change during the observation period with 2324, 2616, and 2160 visits made in February, March, and April by individuals with exactly one annual clinic visit over the prior 12 months.

## Discussion

We found no evidence of a significant drop in overall ambulatory clinic utilization in a rural area of South Africa during the national lockdown for the COVID-19 epidemic. Visits for chronic disease, such as hypertension and diabetes, perinatal care and family planning remained reasonably constant. Notably, child health visits for immunizations and growth monitoring dropped immediately by over 50%, but increased again over time during the lockdown, and neared their pre-lockdown frequency approximately 5 weeks later. We also noted an expected 20% increase in clinic visits for HIV immediately after the lockdown and suspect this might reflect an urgency to collect medications prior to an anticipated interruption in clinic access or medication availability. Our results run counter to our hypothesis, and potentially demonstrate a resilience in the healthcare sector during a period of concern for access to chronic and essential basic health services.

The key demographic population in our study that experienced significant drops in clinic visitation was children. Reassuringly, child health visits appeared to have rebounded during the lockdown and neared (though did not quite return to) their pre-lockdown state, and we did not detect a drop in perinatal care or family planning visits during the lockdown period. However, all-cause visits by children aged 1–5 years old dropped at the initiation of the lockdown period and did not return to pre-lockdown levels by the end of the Level 5 lockdown period. These findings are in keeping with data from the United States, where vaccination rates in children substantially declined after a national emergency was declared in response to the COVID-19 epidemic.^27^ Modeling analyses using Lived Saves Tool (LiST) have suggested that a 15% reduction in maternal and child health coverage could result in over 250,000 additional deaths.^19^ The World Health Organization has also projected significant increases in deaths due to malaria in children under 5 in endemic regions with disruptions in malaria care and insecticide treated bednet distribution.^28^ Previous disease epidemics in sub-Saharan Africa have also been associated with lapses in primary care access, and drops in facility based births and child healthcare access.^20 21 29 30^ Consequently, future work should investigate the impacts of even modest drops in vaccination rates and child health outcomes, to better assess whether the drop we identified resulted in longer term health effects, and whether catch-up vaccination campaigns might help limit the fallout of such interruptions.^31^

Maintaining healthcare access during the epidemic requires a careful balance of primary healthcare provision and protection of vulnerable populations from COVID-19 infection. In other settings, there have been multiple reports of late and severe presentations to care for non-COVID-19 conditions, putatively due to decreased access to care or fear of nosocomial infection at healthcare facilities.^32–34^ By the end of the most stringent Level 5 lockdown period, fewer than 25 cases of COVID-19 infection had been reported in uMkhanyakude District.^35^ Clinics in this district instituted symptom screening at the entryway to clinics, with referral of individuals meeting criteria for persons under investigation to regional COVID-19 testing centres.

The COVID-19 epidemic has also led to calls for decentralized care to minimize exposure for high-risk populations, including those with chronic non-communicable disease, HIV, a history of tuberculosis-related lung disease, and those of older-ages. The lockdown was instituted rapidly in South Africa, before such systems could be put in place. However, an important unanswered question is how such programs will affect access to care and epidemic transmission in high-risk populations, including the elderly and those with immunosuppressing conditions.

Our study should be interpreted within the context of the relatively short period (34 days) of the Level 5 lockdown in South Africa. As a result, we are not yet able to assess longer-term repercussions from disruptions to income or from the epidemic itself, and our results should not be generalized over longer time horizons. It is expected that economic barriers to healthcare utilization will increase as the epidemic’s effects persist over time, including secondary effects from non-pharmaceutical interventions. These effects are likely to fall most heavily on those in the informal economy.^36^ South Africa has taken steps to increase social support to counteract economic disruption from the epidemic and control measures.^37^ Mitigating longer-term consequences will likely require governments and development partners to increase access to employment and other social support services during the epidemic.

Our study had multiple strengths. First, our data collection procedures are led by research staff who remained in place during the lockdown period, so these data are not affected by barriers to data collection (e.g., interruptions in staff transportation or workplace access). This is important, since many routine health information systems could be expected to suffer lapses during external shocks to the healthcare system. Second, our study was able to access data collected across 11 clinical centres within a large HDSS, which provided significant power to detect even small interruptions to health care access. One key potential limitation to our study is that it is predicated on the assumption that there were no other external factors that would have caused interruptions to the health care system on or after 27^th^ March 2020 (e.g., power outage, inclement weather).We are unaware of any such shock and believe this to be a minor risk. Our analysis should also be interpreted within the context of our study area – one with a few dozen reported cases of COVID-19 in a nation with a moderately sized epidemic (approximately 7,000 cases as of early May), but not yet in the depths of a large epidemic with established local transmission.

In summary, we report resilience of the ambulatory health care system during the early COVID-19 epidemic and Level 5 lockdown period in rural South Africa. Future work should establish if these trends are maintained, and particularly monitor access to childcare and immunizations as a result of the trends reported here. Finally, in rural South Africa and similar areas, efforts to prevent nosocomial spread of COVID-19 among high-risk populations through decentralization of non-urgent care will remain a critical area of future study.

## Data Availability

Data from the AHRI HDSS are publicly available upon request to the AHRI research repository which can be made here: https://data.africacentre.ac.za/index.php/auth/login/?destination=

## Transparency declaration

MJS affirms the manuscript is an honest, accurate, and transparent account of the study being reported; that no important aspects of the study have been omitted; and that any discrepancies from the study as originally planned (and, if relevant, registered) have been explained.

## Acknowledgements

We thank the outstanding field staff at the Africa Health Research Institute for their work in conducting the demographic health survey field work, as well as the community and the study participants for their time and efforts. The population HIV surveillance is a node of the South African Population Research Infrastructure Network (SAPRIN) and funded by the South African Department of Science and Innovation and hosted by the South African Medical Research Council. Additional thanks to Zahra Reynolds for her assistance in organizing elements of the manuscript for submission.

